# Dampening of the respiratory cytokine storm is promoted by inhaled budesonide in patients with early COVID-19

**DOI:** 10.1101/2021.10.26.21265512

**Authors:** Jonathan R Baker, Mahdi Mahdi, Dan V Nicolau, Sanjay Ramakrishnan, Peter J Barnes, Jodie L Simpson, Steven P Cass, Richard EK Russell, Louise E Donnelly, Mona Bafadhel

**Affiliations:** National Heart and Lung Institute, Imperial College London, London, United Kingdom; National Institute for Health Research (NIHR) Oxford Biomedical Research Centre (BRC), United Kingdom; Nuffield Department of Medicine, University of Oxford, United Kingdom; UQ Centre for Clinical Research, The University of Queensland, Brisbane, Australia; School of Mathematical Sciences, Queensland University of Technology, Brisbane 4000 Australia; School of Medical and Health Sciences, Edith Cowan University, Perth, Australia; School of Medicine and Public Health, Priority Centre for Healthy Lungs, University of Newcastle, Australia

## Abstract

Vaccinations against SARS-CoV-2 are effective in COVID-19. However, with limited vaccine access, vaccine hesitancy and variant breakthroughs, there is still a need for effective and safe early treatments. Two community-based clinical trials of the inhaled corticosteroid, budesonide, have recently been published showing and improvement in patients with COVID-19 treated early with budesonide^1,2^. To understand mechanistically how budesonide was beneficial, inflammatory mediators were assessed in the nasal mucosa of patients recruited to the Steroids in COVID (STOIC^1^) trial and a cohort of SARS-CoV-2 negative individuals. Here we show that in early COVID-19, elevation in viral response proteins and Th1 and Th2 inflammation occurs. Longitudinal sampling in the natural course of COVID-19 showed persistently high interferon levels and elevated concentrations of the eosinophil chemokine, CCL11. In patients who deteriorate, the initial nasal mucosal signal is characterised by a marked suppression of the early inflammatory response, with reduced concentrations of interferon and inflammatory cytokines, but elevated eosinophil chemokines. Systemic inflammation remained altered in COVID-19 patients, implying that even after symptom resolution, changes in immunological mediators do not resolve. Budesonide treatment decreased IL-33 and IFN-γ, implying a reduction in epithelial damage and dampening of the interferon response. Budesonide treatment also increased CCL17 concentrations, suggesting an improved T-cell response; and significantly alters inflammatory pathways giving further insight into how this treatment can accelerate patient recovery.

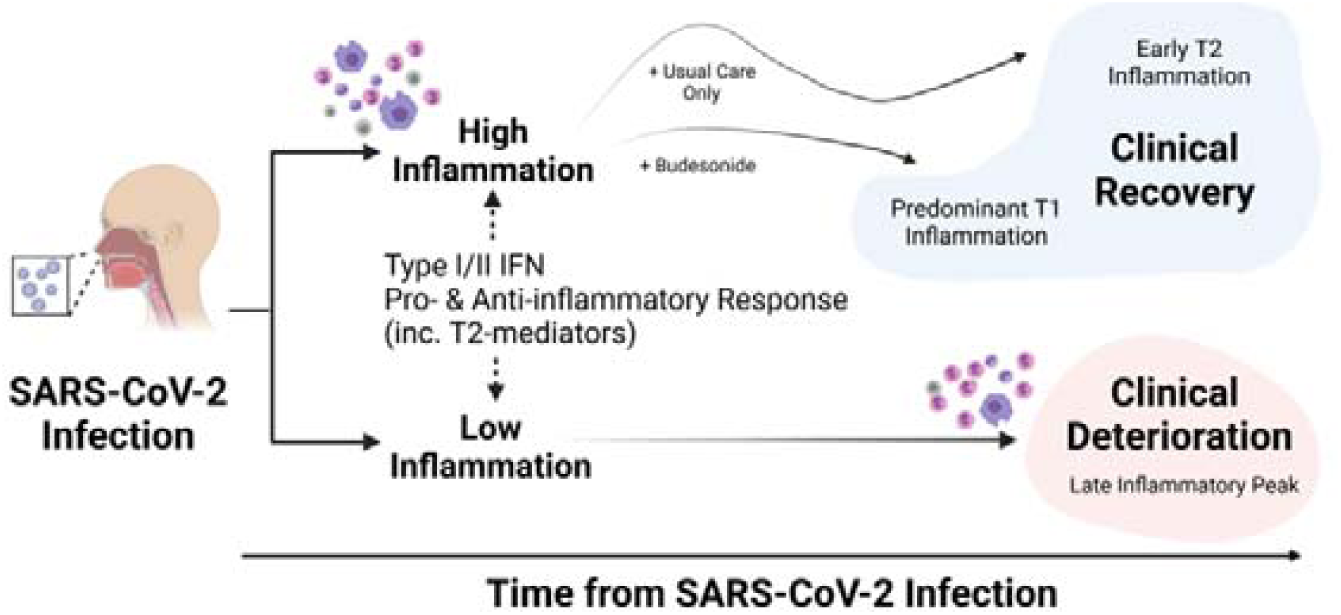

## Introduction

There has been a rapid increase in the understanding of the inflammation that occurs following SARS-CoV-2 infection^3^. Largely this has investigated the immune response in patients with severe infection that required hospitalisation^4^. In the investigation of early COVID-19, studies have focussed on inflammation caused by the systemic immune response^5^ whilst the inflammatory response in the airway of patients remains unknown, in part due to the risk of contagion through aerosolization of the virus. The airway inflammatory response to common respiratory viruses in healthy individuals^6^ and in patients with chronic airways disease has been extensively studied^7,8^. Inhaled corticosteroids (ICS) are often prescribed in patients with asthma and chronic obstructive pulmonary disease (COPD) to reduce the risk of exacerbations which are usually mediated by respiratory viruses^9-11^. During the ongoing COVID-19 pandemic, it has been reported that patients with asthma and COPD are less likely to be hospitalised with severe COVID-19^12,13^. The reasons for this are unclear but in patients with asthma, the use of ICS two weeks prior to hospitalisation for severe COVID-19 was associated with better clinical outcomes^14^. Our recent STOIC study investigated inhaled budesonide, a corticosteroid, as a treatment for early COVID-19 infection with positive findings in improvement in self-reported symptom recovery, with fewer patients experiencing adverse outcomes and less symptom persistence^1^. This finding has now been replicated in a large Phase 3 efficacy trial^2^, but has yet to attract global adoption. The mechanism for how ICS can improve early COVID-19 infection is however currently unknown.

Here, we report the nasal mucosal inflammatory response in patients with early COVID-19 disease and examine the evolution of inflammation in the natural course of COVID-19. We also identify how inflammation in the airway can predict illness severity and investigate the effect of inhaled budesonide upon the respiratory mucosa in early COVID-19 disease. Finally, we show using network analysis how inhaled budesonide can resolve the exaggerated inflammatory response seen in early COVID-19 infection and promote resolution to health.

## Results

The STOIC study recruited 144 participants aged 18-79 (mean 45) years, of which there was 140 available nasal mucosal samples at visit 1 (day 0) and 122 samples at visit 3 (day 14). At day 28 (visit 4), whole blood was collected from 124 participants. The demographics of the healthy volunteers are presented in **supplementary table S1**. The demographics of the STOIC study participants are as previously published^1^.

### The respiratory inflammatory response in early COVID-19 is different to health

Firstly, we examined the inflammatory profile of nasal mucosal fluid in healthy controls (n=20) and early (median duration of symptoms 3 days) COVID-19 infection (n=140). (**Figure 1A**). We found that 14 mediators were elevated in COVID-19 infected patients compared to controls. These were a mixture of Th1 cytokines (IL-2, IL-12, TNF-α, IFN-γ), Th2 cytokines (IL-2, IL-4), interferon/interferon response proteins (IFN-α2a, IFN-β, IFN-γ, CXCL10, CXCL11) and chemokines (CCL3, CCL4, CCL11, CCL13) **(Figure 1B)**. An altered T-cell response with significantly reduced levels of TSLP and CCL17 was also found and reduced CCL2 levels implying impaired monocyte recruitment **(Figure 1B)**. There were also lower levels of VEGF, as has been previously described in severe hospitalised COVID-19 patients^15^ **(Figure 1B)**. Some mediators were unaltered (IL-1β, IL-33, CCL24, CCL26, IL-5, GM-CSF and IL-10), suggesting less activation during early COVID-19 **(Figure 1B)**. CXCL8 was unchanged between health and early COVID-19, but this may reflect assay sensitivity as CXCL8 was repeatedly over the limit of detection (occurred in 35% of healthy control and 45% of early COVID-19 samples).

**Figure 1.**
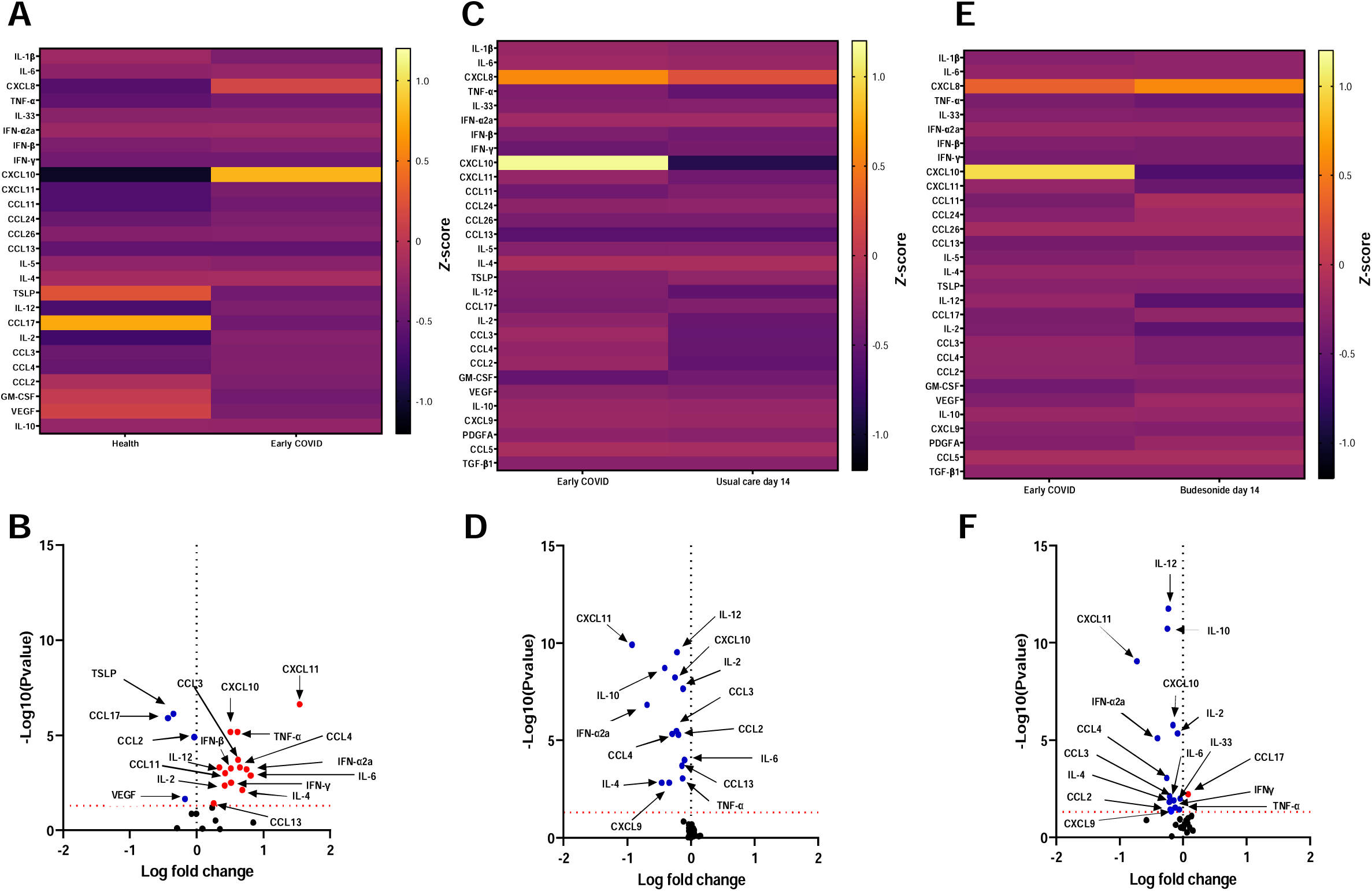
Immunological features of the nasal mucosa in patients with early COVID-19 over time in the STOIC study. A) Heatmap and B) volcano plot of 26 nasal mediators from 20 healthy individuals and 140 patients with community based early COVID-19. C) Heatmap and D) volcano plot of 30 nasal mediators from patients with community based early COVID-19 at visit 1 (Day 0) time point of enrolment into trial and visit 3 (Day 14) in the usual care arm (n=60). E) Heatmap and F) volcano plot of 30 nasal mediators from patients with community based COVID-19 at visit 1 (Day 0) time point of enrolment into trial and visit 3 (Day 14) in the budesonide arm (n=62). Horizontal dotted line on volcano plots depict cut-off for statistical significance, the vertical dotted line represents cut-off point determining whether mediator levels were higher (right, red) or lower (left, blue) for visit 1 samples compared to either healthy controls (unpaired) or paired visit 3 samples. ⍰= upregulated. ⍰= downregulated. Data were analysed using Mann-Whitney t-test or Wilcoxon matched pairs signed rank test.

### Interferon and eosinophil chemotactic proteins remain elevated over the course of early COVID-19

We examined nasal mucosal inflammation during the course of disease (i.e., in the UC study arm, n=60), examining paired samples from this population at visit 1 and visit 3 (**Figure 1C**). Over the course of 14 days, CXCL9, CXCL10, CXCL11, IL-12, IL-10, IL-2, IFNα2a, CCL2, CCL3, CCL4, IL-6, IL-4, CCL13 and TNF-α all significantly reduced. Of the 14 mediators elevated in early COVID-19 compared to healthy controls, 11 of these mediators in the course of early COVID-19 infection decreased over time (CCL3, CCL4, TNF-α, IL-6, CCL13, IL-4, IFN-α2a, CXCL10, CXCL11, IL-2, IL-12). CCL11, IFN-β and IFN-γ did not significantly change between visit 1 and 3 and remained elevated after 14 days compared to health. VEGF, TSLP and CCL17 levels were unchanged over time, remaining lower than health, but CCL2 and IL-10 levels significantly reduced **(Figure 1C**). IL-1β, CXCL8, IL-33, CCL24, CCL26, IL-5, GM-CSF did not show any changes between the two study visits over time. In the nasal mucosa of participants in the study, we also assessed a further 4 mediators, of which PDGFA, CCL5 and TGF-β1 were unchanged over time; whilst CXCL9 levels reduced (**Figure 1D**).

### Inhaled budesonide leads to reduction of Type 2 cytokines over the course of early COVID-19

The STOIC study showed that inhaled budesonide treatment, taken for a mean duration of 7 days, reduced the number of COVID-19 patients who deteriorate and reduces the time to clinical recovery^1^. To investigate the potential mechanisms behind this finding we examined the concentrations of 30 mediators in paired visit 1 and visit 3 samples in the inhaled budesonide arm of the STOIC trial (n=62, **Figure 1E**). Of the mediators examined, those that were different to the natural course of early COVID-19 disease (as described above from the UC arm), IL-33 and IFN-γ were significantly reduced, CCL17 concentrations were significantly increased, and CCL13 was no longer significantly reduced between the two visits (**Figure 1F**). The differences between UC and BUD, showed that IL-33 was significantly reduced and CCL17 significantly elevated in the BUD arm (**supplementary figure S2**). As all visit 1 samples were taken prior to treatment, we also compared both the UC study arm and BUD study arm visit 3 samples to all visit 1 samples to assess the return to health. We found that following treatment with inhaled budesonide, CCL5 was significantly reduced, and GM-CSF was not reduced; whilst concentrations of IL-2 and IL-4 were sustained, despite falling in the UC study arm **(supplementary figure S3**). Changes in the levels of these mediators may therefore help to explain differences in the recovery of these patients and play a part in the disease course of COVID-19.

### SARS-CoV-2 viral load correlation with nasal mucosal inflammation differs from systemic inflammation

As previously reported viral load correlates with type I interferon expression and inflammatory markers and chemokines in the blood^16^. We thus examined whether there was a similar correlation with the nasal mucosa. SARS-CoV-2 viral load and nasal mucosal mediator measurements were available in 122 paired samples at study entry (day 0). We found that IFN-α2a, CXCL10, CXCL11, IL-12, CCL2 and IL-6 correlated with viral load **(supplementary table S2**). These show a different pattern of markers, with some overlap, to those measured in serum^16^ and may reflect differences between local and systemic inflammation, perhaps related to the number of viral particles.

### Despite clinical recovery, nasal mucosal inflammation does not completely return to normal

To assess whether the inflammation associated with SARS-CoV-2 infection declined over time to levels in health, we compared our healthy cohort to the visit 3 samples of the UC and BUD study arms **(Figure 2A)**. In both UC and BUD study arms, compared to healthy controls, IL-2, CCL11, IL-4, TNF-α, IFN-β, IFN-γ and CXCL11 all remained significantly elevated, whilst CCL3, CCL4, IL-6, CCL13, IFN-α2a, CXCL11 and IL-12 reduced back to comparable healthy concentrations (**Figure 2B**). CCL2 and TSLP remained significantly lower than health irrespective of the study arm. Even though the concentration of CCL17 was increased in the BUD study arm over time, these did not return to expected healthy concentrations. Interestingly, IL-10 significantly decreased over time in the UC study arm but not in the BUD study arm, suggesting a sustained anti-inflammatory response in patients treated with inhaled budesonide (**Figure 2B**). It should be noted that the time to self-reported clinical recovery in the study was a mean of 9 days, suggesting that although certain mediators reduce during the disease, multiple mediators remain increased within the nasal mucosa and significantly past the time-point of clinical recovery.

**Figure 2.**
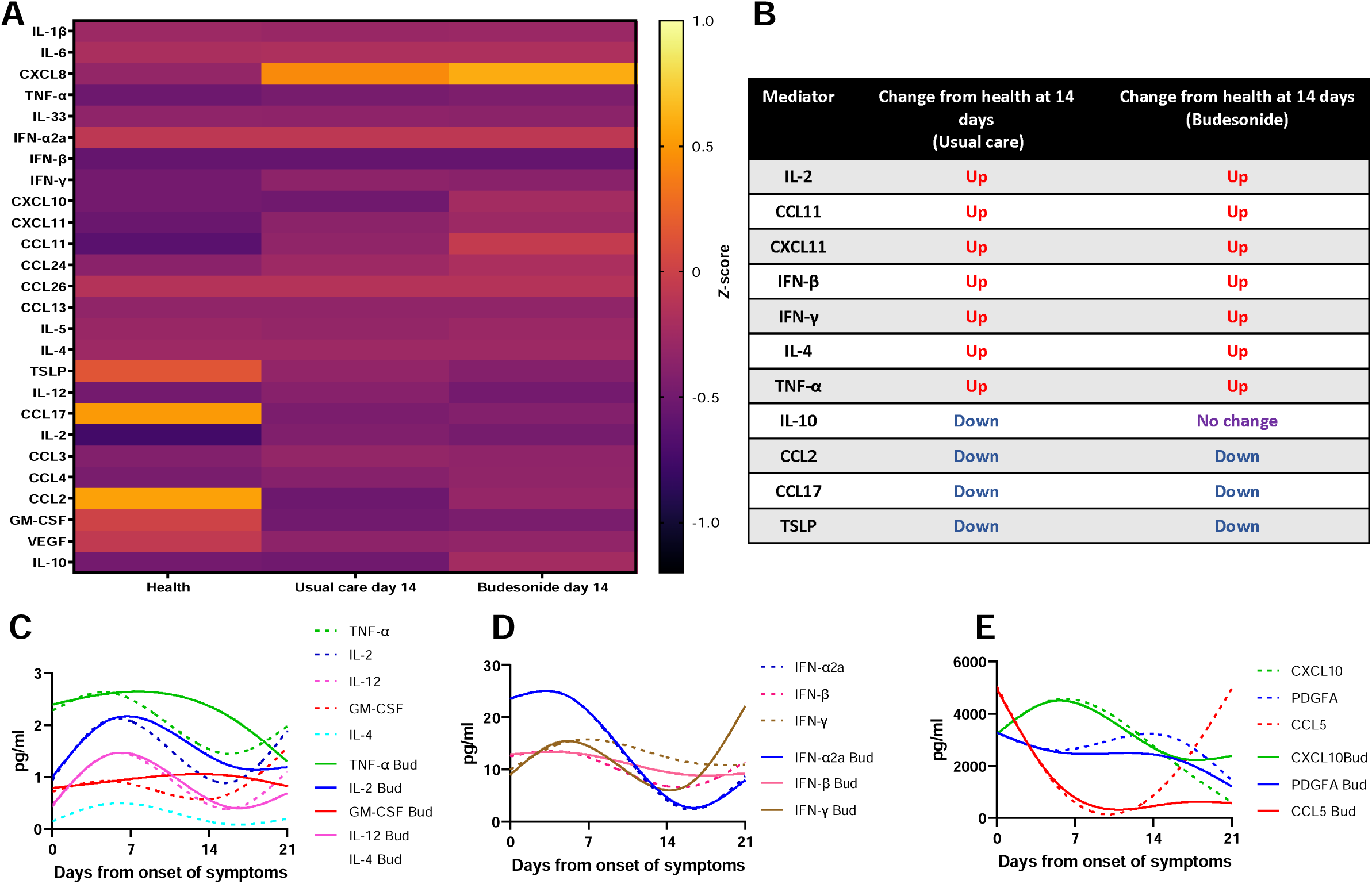
Temporal changes in mediator levels in usual care and budesonide arm. A) Heatmap of 26 nasal mediators from 20 healthy individuals compared to samples at day 14 post recruitment (visit 3) from usual care study arm (n=60) and budesonide study arm (n=62). B) List of significantly altered mediators compared from health at day 14 time point in both usual care and budesonide treated arm. C-E) Longitudinal analysis of mediator profiles in the usual care and budesonide arm displayed as representative best fit curves by smoothed spline analysis.

### A bi-phasic peak of inflammation is seen in the course of COVID-19, which is supressed by inhaled budesonide

Spline best-fit curve analysis was performed to assess kinetic inflammatory changes over the disease course of early COVID-19 infection (UC study arm) and with ICS intervention (BUD study arm). The spline analysis considers the timing of the first symptom and gives an indication of peak of inflammation and resolution. In general, peak inflammation occurred early in the disease course. Additionally, IL-4, IL-12, IL-2, CXCL10, PDGFA and IFN-α2a kinetic changes were similar between natural course of early COVID-19 and inhaled budesonide. However, inhaled budesonide supressed kinetic inflammation and in particular a secondary increase in inflammation beyond day 15 (as seen with TNF-α, GM-CSF and CCL5). Interestingly, IFN-γ increased following inhaled budesonide at day 15 following symptom onset **(Figure 2C - 2E and supplementary figure S4)**.

### The early nasal mucosal inflammatory pattern can predict patients that deteriorate following COVID-19 infection

Eleven participants met the primary outcome for deterioration in the clinical trial (defined as needing urgent care assessment, emergency care consultation and/or hospitalisation), with 70% needing oxygen treatment. Spline mediator analysis was performed to investigate the kinetics of nasal mucosal inflammation from the first 7 days of symptom onset. Specifically, IL-2, IL-12, IL-4, GM-CSF, IL-10, IL-5, IFN-γ did not increased, whilst CXCL9, CXCL10, TNF-α and IFN-β showed peaks much higher and earlier than samples from participants without clinical deterioration following COVID-19 infection **(supplementary figure S5)**. Comparison of mediators from healthy controls and participants that met the primary outcome showed that CCL3 TNF-α, IFN-α2a, IFN-β, CXCL10, CXCL11, CCL11 and CCL24 were all significantly increased. CCL3, TSLP, CCL17, IL-10, GM-CSF and IL-33 were significantly reduced (**Figure 3A**). To try and understand why these participants met the primary outcome, we examined mediator concentrations in these participants against health (**Figure 3B**). We found that nasal mucosal inflammation in participants who reached the primary outcome, did not respond to SARS-CoV-2 infection in the same manner. There was no increase in CCL4, IL-12, IFN-γ, IL-4, CCL13, IL-2, and IL-6, suggesting an impaired inflammatory response. Additionally, reduction in IL-10, IL-33 and GM-CSF occurred in this group compared to no change in the early COVID-19 cohort. VEGF did not reduce in those who met the primary outcome opposed to those with early COVID-19. Interestingly, CCL24 levels were increased in participants who met the primary outcome, confirming that a high eosinophilic phenotype is associated with severe disease.

**Figure 3.**
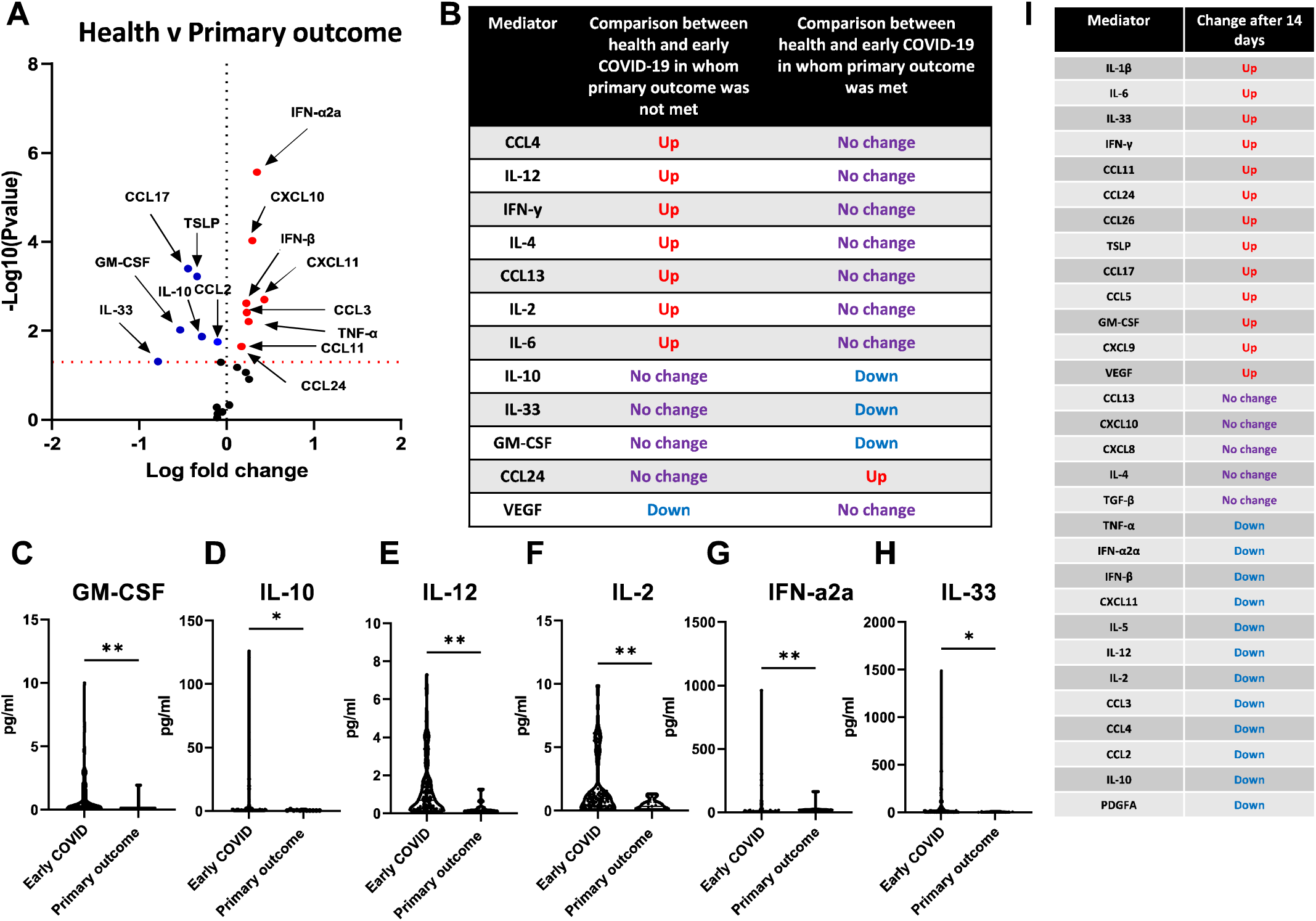
Alterations in nasal mucosal inflammation in whom early COVID-19 clinically worsens. A) Volcano plot of 26 nasal mediators from 20 healthy individuals and 11 patients in who the primary outcome of the study was met; ⍰ = significantly upregulated. ⍰= significantly downregulated. B) Comparison table of significantly altered mediators from nasal samples comparing health (n=20), early COVID-19 (n=129) without deterioration and COVID-19 with deterioration (n=11). C-H) Significantly altered mediators at visit 1 from patients with early COVID-19 (n=129) and those who met the primary outcome (n=11). I) Change in levels of 30 mediators from visit 1 (day 0) to visit 3 (day 14) of 1 patient who required critical care respiratory support **(see supplement S6 for heatmap)**. Data were analysed using Mann-Whitney t-test, *= p<0.05, **= p<0.01.

Comparison of mediators in participants with COVID-19 and participants with COVID-19 who clinically deteriorated was performed to understand any aberrant immune response during the early part of SARS-CoV-2 infection. Interestingly, all mediators that were significantly different (GM-CSF, IL-10, IL-12, IL-2, IFN-α2a and IL-33) were downregulated in worsening disease suggesting at the early stages of the disease a blunted response to infection occurs in patients who then deteriorate (**Figure 3C-H**). In one participant, needing critical care support (UC study arm), longitudinal nasal sampling was available giving insight into course of the disease in a more severe patient. Interestingly, the interferon response proteins, CCL5, CXCL9 and CXCL10 remained elevated and CXCL11 fell. Concentrations of the inflammatory markers IL-1β, IL-33 and IL-6 also increased over time in this participant (**Figure 3I, supplementary figure 6)**.

### There is persistent systemic inflammation following COVID-19 infection, but this is attenuated by inhaled budesonide

Serum inflammatory mediators up to 35 days after initial infection remained elevated compared to healthy controls (**Figure 4A-F, supplementary figure 7)**. This included higher concentrations of TNF-α, IL-8 and the family of eosinophil chemotactic proteins (CCL11, CCL24, CCL26) and may be a driver for long COVID. This systemic inflammation was supressed to a greater degree by inhaled budesonide, for CCL11, TNF-α and IL-33.

**Figure 4.**
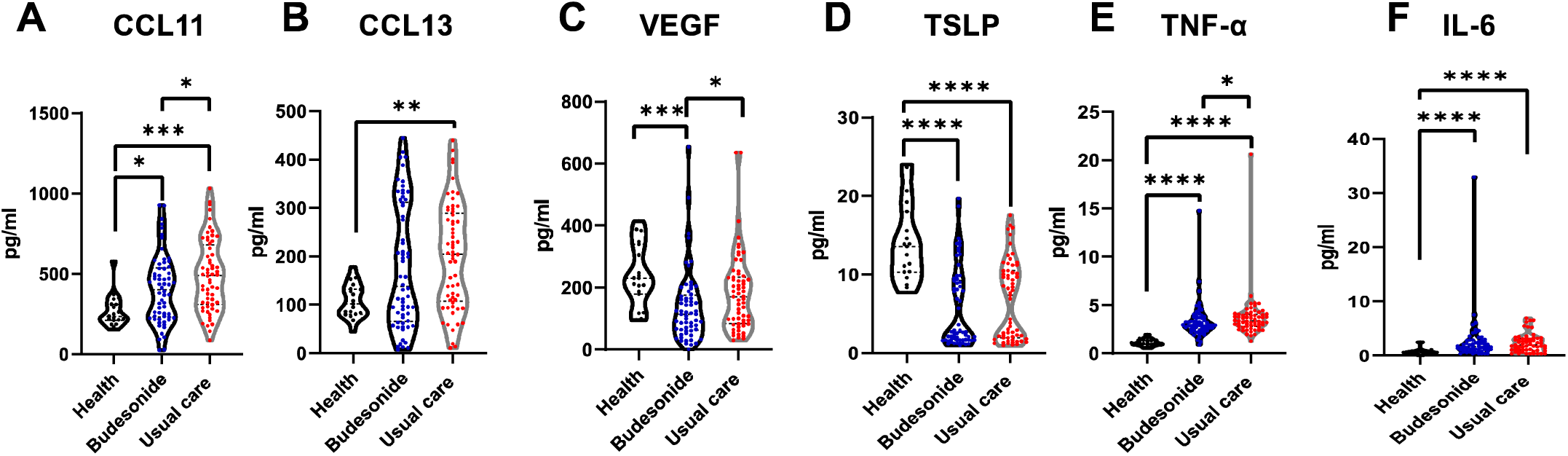
Persistence of systemic inflammation following 28-35 days of COVID-19 infection in the community, is dampened by inhaled budesonide. A-F) Violin plots comparing some mediator levels in the serum of healthy individuals (n=19), those in the usual care arm (n=60) and budesonide arm (n=62) of the study at 28-35 days following COVID-19 infection (**see supplement S7 for further results**). Data were analysed by Kruskal-Wallis with post-hoc Dunn’s test. * p<0.05, ** p<0.01, ***p<0.001, ****p<0.0001.

### Effect of inhaled budesonide treatment on mediator expression following COVID-19 demonstrated normalised T2 airway and circulatory response

Network analysis of inflammatory mediators in the nasal mucosa and serum were performed (**Figure 5**). A rapid onset anti-inflammatory response (IL-10) and T2 inflammatory (CCL11 and CCL24) response with co-existent inflammation (IL-6 and CXCL8) can be seen in early COVID-19 infection (**Figure 5A-1**). Two weeks after initial infection, the natural course of COVID-19 infection shows persistent nasal mucosal inflammation with an exaggerated T2 inflammatory response (CCL11 and IL-5) and a sustained pro-inflammatory response (**Figure 5A-2)**. Network analysis demonstrated that there was 72% overlap between the inflammatory response at an early timepoint in COVID-19 infection and at the later timepoint, indicating persistent inflammation of similar pathways. Following inhaled budesonide treatment, the nasal mucosal response is different with an increased anti-inflammatory response (IL-10) and alarmins (IL-33 and TSLP), but a reduced T2 inflammatory response suggesting normalisation of the anti-viral immune response with suppression of the T2 hyper-inflammatory response we have seen following COVID-19 infection. The overlap between the inflammatory response at an early timepoint in COVID-19 infection and at the later timepoint was only 27% in the BUD study arm. Network analysis of serum mediators performed between 28 and 35 days after initial infection in the UC study arm shows persistent IFN-α2a, CCL11 and CCL2 (**Figure 5B-1**). There was only a 30% overlap with the serum of participants that had been treated with inhaled budesonide (**Figure 5B-2**). These findings suggests that inhaled budesonide modulates the inflammatory pathways in the upper respiratory tract and circulation following COVID-19 infection.

**Figure 5.**
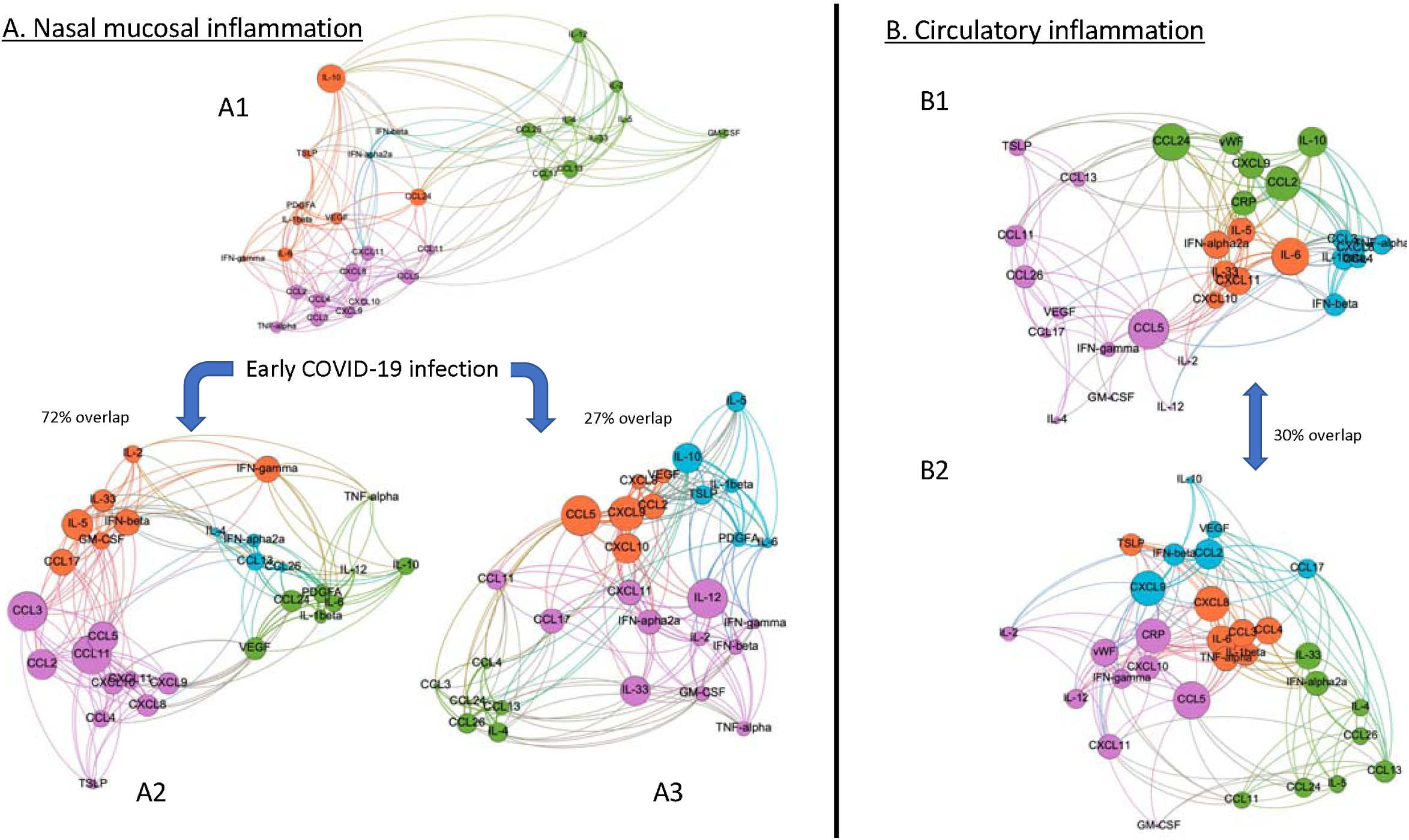
Network analysis of mediator correlation data. **Panel A:** Modularity maximisation and community detection for eigenvalue-based noise-cleaned nasal mucosal mediator data. **Panel A1** corresponds to networks at initial COVID-19 infection; **Panel A2** corresponds to networks after 14 days of initial COVID-19 infection; and **Panel A3** corresponds to networks after treatment with inhaled budesonide. Node size indicates eigenvalue centrality and arcs indicate non-zero noise-cleaned correlation. In each network there are four modules, of roughly equal size. Panel A1 and A2 are similar with 72% overlap indicating persistent relationship of inflammation over time. Panel A1 and A3 have 27% overlap (which would be expected by a chance reassignment of nodes to the four modules) indicating that treatment with inhaled budesonide changes the inflammatory networks entirely. **Panel B**: Modularity maximisation and community detection for eigenvalue-based noise-cleaned serum data in participants from 28 to 35 days following COVID-19 infection. **Panel B1** is network analysis of serum from participants in the usual care arm. **Panel B2** is the network analysis of serum from participants in the budesonide arm. The two networks are entirely different in terms of membership with 30% overlap (equivalent to a random node reassignment to modules). This analysis is also consistent with a model in which budesonide treatment dramatically rearranges the mediator pathways in serum.

## Discussion

In this study, we examined the inflammatory effect of SARS-CoV-2 upon the upper airway at a very early timepoint during the course of COVID-19 infection^1^ and followed patients over the evolutionary course of COVID-19 infection. Our study provides a unique mechanistic insight into the host respiratory immune and inflammatory response to early COVID-19 infection and following a therapeutic intervention with benefit. Notably, studies thus far have examined only a small number of airway samples from severe and hospitalised patients, late in the disease course^17,18^ or measured the immune response in the circulation^19^. Moreover, these studies focus on transcriptional analysis which may not fully recapitulate the protein environment. In contrast this study is the first to examine protein levels in the upper respiratory tract of early COVID-19; thus providing a unique mechanistic insight into the host respiratory immune and inflammatory response to early COVID-19 infection and subsequent therapeutic intervention with benefit.

Our study has shown, unlike previously assumptions^20^, that there is a vigorous and early immune response in the upper airway in patients who develop COVID-19 infection. This response, in comparison to healthy controls, is an early combined Type I and Type II interferon response, pro-inflammatory response and anti-inflammatory response. Interestingly, early in COVID-19 there is also a T2 immune response, mediated mainly by CCL11 and IL-4. This early COVID-19 innate cytokine immune response will lead to recruitment of eosinophils, NK cells and macrophages and is an immune response similar to that which is seen with other respiratory viruses^21^.

We have also shown that in patients in whom there was a clinical deterioration following SARS-CoV-2 infection, the initial inflammatory response in the airway is blunted with an impaired inflammatory response and reduced concentrations of IL-2, IL-33, IL-12 and IFN-α2a. A blunted circulatory interferon response has been previously shown to be a feature of severe COVID-19 infection^22^. As our sampling was in early COVID-19 infection, these findings suggest that an early impaired interferon response to COVID-19 can highlight an important aspect of diagnosis and treatment; and is a potential biomarker to predicting patients at risk of deterioration who warrant further early treatment. Identification of patients early and at risk of deterioration will markedly improve clinical care for patients.

Interestingly, CCL24 was the only mediator increased in patients who deteriorated following COVID-19 infection. The CC subfamily of eosinophil chemotactic proteins (CCL11, CCL24 and CCL26), along with IL-5, GM-CSF and CCL5 are responsible for lung recruitment and activation of eosinophils^23^. Recruitment of eosinophils to the lung, is the most likely explanation for the reduced circulatory number of eosinophils reported as part of the clinical picture of severe COVID-19 infection^24^. Furthermore, inhaled corticosteroids are recognised to be therapeutically effective in patients with an eosinophilic phenotype of airways disease^25^. The elevation of CCL24 in patients that subsequently deteriorated in this study validates that the high eosinophilic phenotype, albeit measured in the plasma previously is associated with severe disease, as previously described^16^.

Our study is also the first to demonstrate that there is ongoing inflammation in the circulation, over 4 weeks after SARS-CoV-2 infection, with elevated levels of IL-2, IL-6, CXCL8, CCL13, TNF-α and interestingly CCL11, CCL24 and CCL26. Persistent symptoms following COVID-19 infection has been recently been identified as an emerging problem^26^ and our study may implicate how this may be underlined by persistent inflammation. To note, the airway and peripheral blood immune profile in severe COVID-19 infection is different^27^. It is therefore plausible that the identification of inflammatory mediators in the circulation is delayed compared to assessing samples proximal to the site of infection. The STOIC study showed that persistence of symptoms was significantly lower in patients that were taking inhaled budesonide suggesting that early inhaled corticosteroid treatment can improve symptoms of COVID-19 and may help to prevent the effects of long COVID. We found that TNF-α and CCL11 were significantly lower in the systemic circulation in patients that had been treated with inhaled budesonide compared to usual care, suggesting that these mediators could be key for future clinical targets. Patients with long COVID commonly report symptoms of breathlessness^28^, whilst asthma has been found to be the only pre-existing comorbidity with an independent association of long COVID^28^. Whether there is a pre-existing host or exaggeration of the T2 phenotype that leads to long COVID warrants further investigation.

Due to the nature of the STOIC study design^1^, we were able to examine the kinetics of inflammation amongst patients with early COVID-19 and in patients that received inhaled budesonide as a treatment intervention. We found that in early COVID-19 there is down-regulation of inflammation, but this does not return to levels seen in health. Examination of the mediator temporal relationship showed that there was often a bi-phasic peak of inflammatory mediators, which is staggered and persists over time. Importantly however, we found that in patients taking inhaled budesonide, the inflammatory peak was attenuated for TNF-α, GM-CSF, and CCL5 and promoted for IFN-γ. IFN-γ is an important promoter of anti-viral immunity^29^ playing a role in the inhibition of viral replication in cells^30^, suggesting another mechanism for the efficacy of ICS. Following inhaled budesonide treatment, the nasal mucosal response is different with an anti-inflammatory response (IL-10), an increase in alarmins (IL-33 and TSLP) and a reduced T2 inflammatory response which suggests the promotion of a normal anti-viral and suppression of the exaggerated T2 inflammatory response.

The initial observation of a significant under-representation of patients with obstructive lung disease (asthma and COPD) with severe COVID-19 infection, with the knowledge that ICS treatment is routinely used to prevent viral exacerbations was the founding hypothesis for the STOIC trial^1^. The respiratory epithelium, especially the nasal mucosa exhibits the most abundant expression of ACE2 receptors important for SARS-CoV-2 binding^31^. ICS have been shown to reduce ACE2 receptor expression *in-vitro*^32^ and *in-vivo*^33^, plus have previously been shown to inhibit viral replication *in-vitro* and reduce inflammation following rhinovirus infection^34^. The potential therapeutic effect of ICS in other respiratory virus infections have previously been explored^35^. ADAM17 is a necessary protease for ACE2 receptor availability and also mediates the release of IL-6 and TNF-α^36^. ADAM17 gene expression is reduced following ICS therapy^33^. ICS reduce inflammation by decreasing the transcription of inflammatory cytokines including IL-4, IL-5 and GM-CSF and the chemokines, CCL11 and CCL5^35^. These pathways form part of a Th2 inflammatory response that we have seen in our study of early COVID-19, and particularly in patients that deteriorate. Our findings suggest that we now have a therapeutic mechanism of how inhaled corticosteroids are an effective treatment in early COVID-19 infection.

## Methods

### Study design and participants

STOIC was a randomised, open-label, parallel group, phase 2 clinical intervention trial. Participants aged 18 and over with early COVID-19 symptoms (defined as new onset of cough and/or fever and/or anosmia for less than 7 days) were randomised to receive usual care (UC), namely as required anti-pyretics, or inhaled budesonide (BUD) at a dose of 800μg twice a day plus usual care (**see supplement figure S1**). Participants were seen at home at randomisation day 0 (visit 1), day 7 (visit 2), and day 14 (visit 3) by a research nurse to obtain consent, provide inhalers and collect nasal absorption samples and nasopharyngeal swabs which were self-performed. At day 28 (visit 4), whole blood was collected. Full details of study design, research protocols and clinical trial results are published^1^. The study was approved by Fulham London Research Ethics Committee (20/HRA/2531) and the National Health Research Authority and was sponsored by the University of Oxford. All participants provided written informed consent.

### Healthy control participants

Healthy controls were adults aged 18 and over, without any known history of lung disease or symptoms (including negative COVID-19 antigen tests) of COVID-19, recruited from a long-term observational data collection study at the University of Oxford (Ethics Ref 18/SC/0361).

### Nasal mucosal sampling

Nasal mucosal sampling was self-performed by all participants as previously described^37^. Briefly, samples of the nasal mucosal lining fluid were collected by placing a Nasosorption™ FX·I device (Hunt Developments UK Ltd) consisting of a synthetic absorptive matrix strip against the inferior turbinate for a duration of 1 minute. The sample was then eluted in 500 μL of PBS, 1% bovine serum albumin (BSA) (w/v), 1% Triton X-100 (v/v), and 0.05% sodium azide (w/v) (Sigma-Aldrich, UK) and stored at -80°C.

### Isolation of serum

Whole blood was collected into an SST tube (Fisher Scientific, Leicestershire, UK) and allowed to clot for 60 min at room temperature. The serum sample was then centrifuged at 3000 x g for 10 min at room temperature. Serum samples were then aliquoted and stored at -80°C.

### Virus extraction and quantification

RNA was extracted using the QIAamp Viral RNA Mini Kit following manufacturer’s instructions. Virologic testing for SARS-CoV-2 infection was performed by quantitative real-time RT-PCR (RT-qPCR). Briefly, 420 µl of sample was extracted and eluted into 40 μl Buffer AVE. 10 µl of eluted RNA was assayed using the Taqman fast virus 1-step master mix (ThermoFischer Scientific, Loughborough, UK), utilising oligonucleotide primers (600 nM forward and 800 nM reverse per reaction) and fluorescent conjugated probes (two probes 100 nM each) (Eurofins Genomics, Wolverhampton, UK) for the detection of the viral RNase P gene (RdRP) gene region of SARS-CoV-19. Quantification of virus RNA copies were generated using a standard curve using nCoV-WHO-Control plasmid of known concentration (Eurofins Genomics, Wolverhampton, UK).

### Immunoassays

Immunoassays of mediators were selected to represent putative mechanistic inflammatory pathways associated with respiratory virus infections and thus COVID-19^15,16,38^. Granulocyte macrophage-colony stimulating factor (GM-CSF), vascular endothelial growth factor (VEGF), tumour necrosis factor (TNF)-α, interferon (IFN)-α2a, IFN-β, IFN-γ, IL-33, thymic stromal lymphopoietin (TSLP), and interleukin (IL-)--1β, IL-2, IL-4, IL-5, IL-6, CXCL8, IL-10, IL-12, CXCL10, CXCL11, CCL2, CCL3,

CCL4, CCL13, CCL17, CCL11, CCL24 and CCL26 were quantified using MSD (Meso Scale Diagnostics, Rockville, Maryland, USA) 10 spot U-Plex plates. Transforming growth factor (TGF)-β1, platelet derived growth factor (PDGF)-A, CXCL9, CCL5, von-Willebrand factor (vWF) and high-sensitivity C reactive protein (hsCRP) were measured by individual V-Plex plates, on a SQ120 Quickplex instrument (Meso Scale Diagnostics, Rockville, Maryland, USA). All values at or below the lower limit of detection (LLOD) or upper limit of detection (ULOD) were replaced by the LLOD or ULOD value as suggested by the assay parameters. Nasal mucosal mediators that were consistently around the ULOD were CXCL10 (6000 pg/ml) and CXCL8 (2100 pg/ml) whilst IFN-α2a (4 pg/ml), CCL26 (7.3 pg/ml), IL-4 (0.08 pg/ml) and TGF-β1 (9.1 pg/ml) were around the LLOD.

### Statistical analysis

Statistical analyses used GraphPad Prism v9.0.0 (GraphPad, La Jolla, California, USA). Spline analysis was calculated by correlating at mediator levels and time from symptom onset as previously described^39^. Z-scores were calculated using the mean and standard deviation of all values dependent on analyses. Volcano plots were generated by calculating the -LOG10 of the P value change between healthy control and all visit 1 COVID-19 patients (un-paired analysis by Mann-Whitney) or visit 1 from the usual care study arm (UC) and budesonide study arm (BUD) against their paired visit 3 sample (paired analysis by Wilcoxon signed-rank test). Serum samples were analysed by Kruskall-Wallis with post hoc Dunn’s post-hoc multiple comparison test. Network analysis and visualisation software was developed in-house (code available on request) and is described below. For image rendering the Gephi package v0.9.2 (Gephi.org. The Open Graph Viz Platform) was used. P values <0.05 was considered statistically significant.

### Network analysis of inflammatory mediators

To further elucidate the patterns of interactions between inflammatory mediators in COVID-19 both with and without inhaled budesonide treatment, we performed a mathematical analysis of the networks of mediators in various subsets of our data.

First, we computed correlation coefficients between mediator values in the nasal data across patients. For each of M mediators, we have values for n patients in either the BUD or UC arms. Regarding each patient index as an independent variable and each mediator value as a dependent variable, we can compute mediator-mediator expression correlation values, yielding an m x m matrix, with 1s on the diagonal.

This matrix represents a network with *m* mediator nodes and whose entries correspond to the strength of mediator co-expression. In principle this network can be analysed as-is. However, it is known that correlations between data series containing measurement error contain as a result a large component of ‘noise’. In particular, the eigenvalues of a rectangular matrix whose entries are random follow the Marchenko-Pastur distribution^40^. Therefore, those eigenvalues (and their associated eigenvectors) for an *m* x *m* correlation matrix falling within this distribution correspond to noise in the data and should be removed from the matrix. Specifically, the eigenvalues falling between the bounds

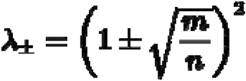

correspond to noise. In the types of correlation matrices analysed here (where *n* is not much larger than *m*), most eigenvalues fall between these bounds (i.e., most of a correlation matrix is actually noise), confirming that analysing the original correlation networks directly would yield highly unreliable findings. By setting the noise eigenvalues to zero and reconstituting the matrix from the remaining non-zero (signal-associated) eigenvalues and their eigenvectors, we get an “eigen-cleaned” matrix, which can be used to perform network analysis on the co-expression of mediators (code available on request). We performed this procedure for the following data sets:

1. Early COVID-19 infection at study enrolment (visit 1, baseline) for nasal mucosal inflammatory data
2. BUD and UC arm analysis at visit 3 (14 days after randomisation) for nasal mucosal inflammatory data
3. Serum mediator data for participants 28-35days after COVID-19 infection in the BUD and UC study arms respectively

This procedure results in 6 mediator-mediator co-expression networks. We split the analysis into the nasal and serum components. In the former case, we can look at how the visit 3 UC and BUD networks differed from the early COVID-19 baseline network, as well as from each other. In the latter case, we can compare the UC and BUD visit 3 networks with each other as well as with the healthy control network. We used modularity maximisation (code available on request) to detect communities (modules) within each of these networks. We chose the well-known Louvain algorithm for this analysis^41^ due to its robustness. Remarkably, in each of these networks, the algorithm detects four sub-modules that are highly internally connected and relatively unconnected from the other modules, of approximately equal size (4-8 mediators) confirming that there is substantial structure in mediator-mediator co-expression. However, the mediator membership assignment to the modules in each case varies substantially and, in particular, is altered by treatment with budesonide versus usual care.

## Data Availability

The datasets generated during and analysed during the current study are available from the corresponding author on reasonable request

## Acknowledgements

The study was funded by the Oxford NIHR Biomedical Research Centre and AstraZeneca (Gothenburg Sweden). The funders had no role in study design, data collection, data analysis or the decision to publish. The views expressed are those of the authors and not necessarily those of the NHS, the NIHR or the Department of Health.

## Author contributions

JRB, MM and SR were responsible for all sample collection, processing and analysis. JRB, DVN, PJB, JLS, SPC, REKR, LED and MB were responsible for data interpretation critical revision of the work. All authors contributed to this manuscript and were involved in the writing and approved the final submission.

## Competing interest declaration

JRB, SPC, MM, DVN, and JSL have no competing interests to declare. SR reports grants and non-financial support from Oxford Respiratory NIHR BRC during the conduct of the study; non-financial support from AstraZeneca, personal fees from Australian Government Research Training Program outside the submitted work. LED reports grants from AstraZeneca, from Boehringer-Ingelheim, outside the submitted work. PJB reports grants and personal fees from AstraZeneca, grants and personal fees from Boehringer Ingelheim, personal fees from Teva, personal fees from Covis during the conduct of the study. REKR reports grants from AstraZeneca, personal fees from Boehringer Ingelheim, personal fees from Chiesi UK, personal fees from Glaxo-SmithKline during the conduct of the study. MB reports grants from AstraZeneca, personal fees from AstraZeneca, Chiesi, GSK, other from Albus Health, ProAxsis, outside the submitted work.

## Extended Data Supplementary Figures

**Supplementary figure S1.**
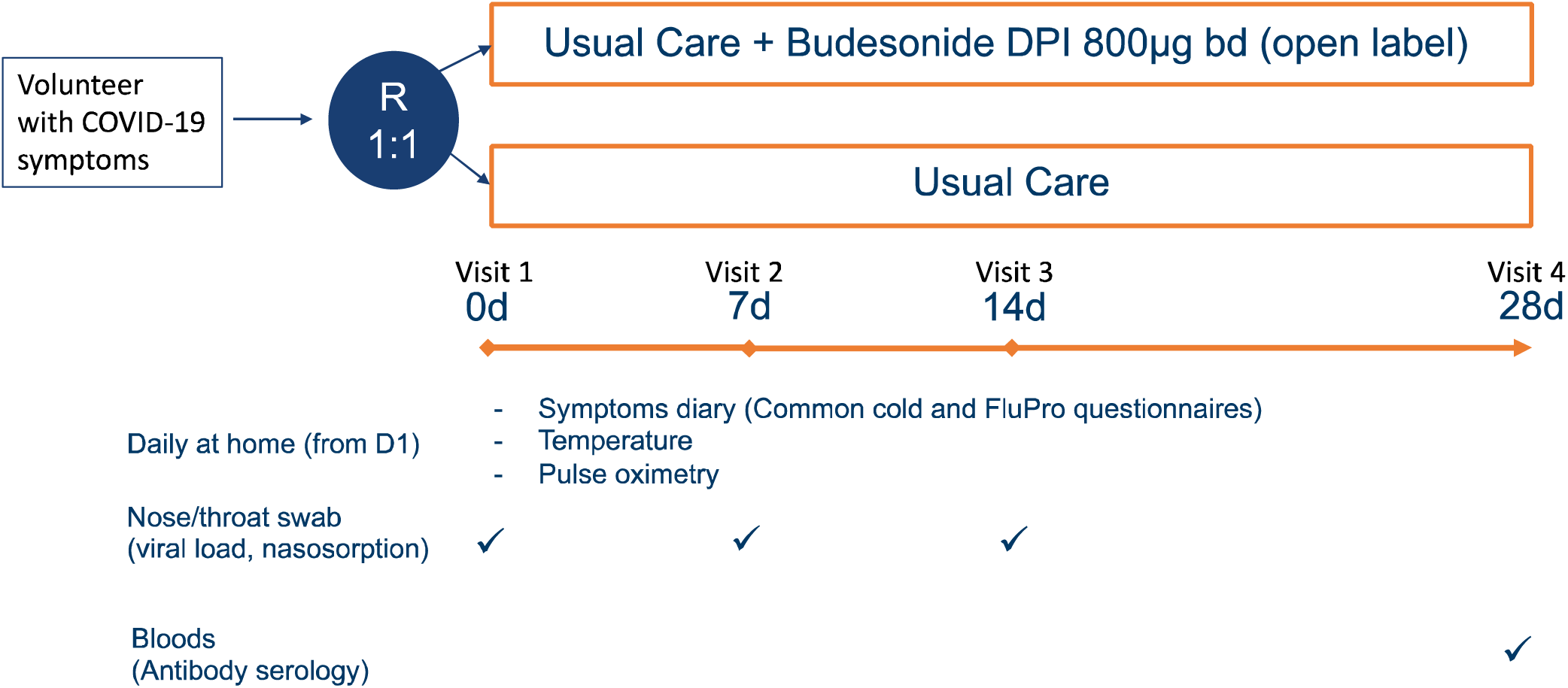
Study design schematic for the STOIC study. DPI Dry powdered inhaler; bd (twice per day); d days

**Supplementary figure S2.**
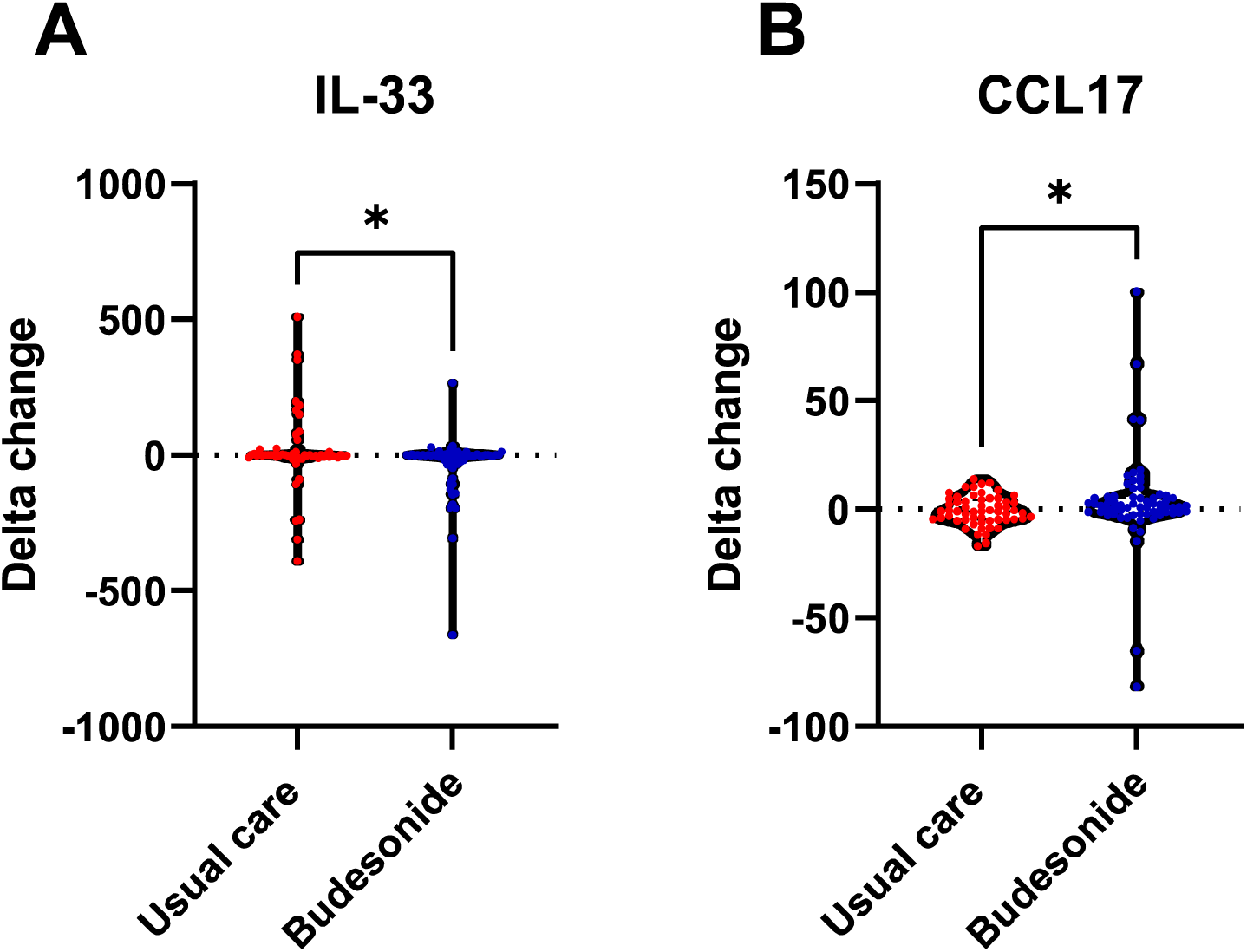
Delta changes between Usual care and Budesonide arm. Significant Delta changes in protein levels between visit 1 and visit 3 between usual care arm and budesonide treated arm. Delta changes in A) IL-33 and b) CCL17 comparing the visit 3 samples from usual care (n=60) and budesonide (n=62). Data were analysed using Mann-Whitney t-test. *P<0.05

**Supplementary figure S3.**
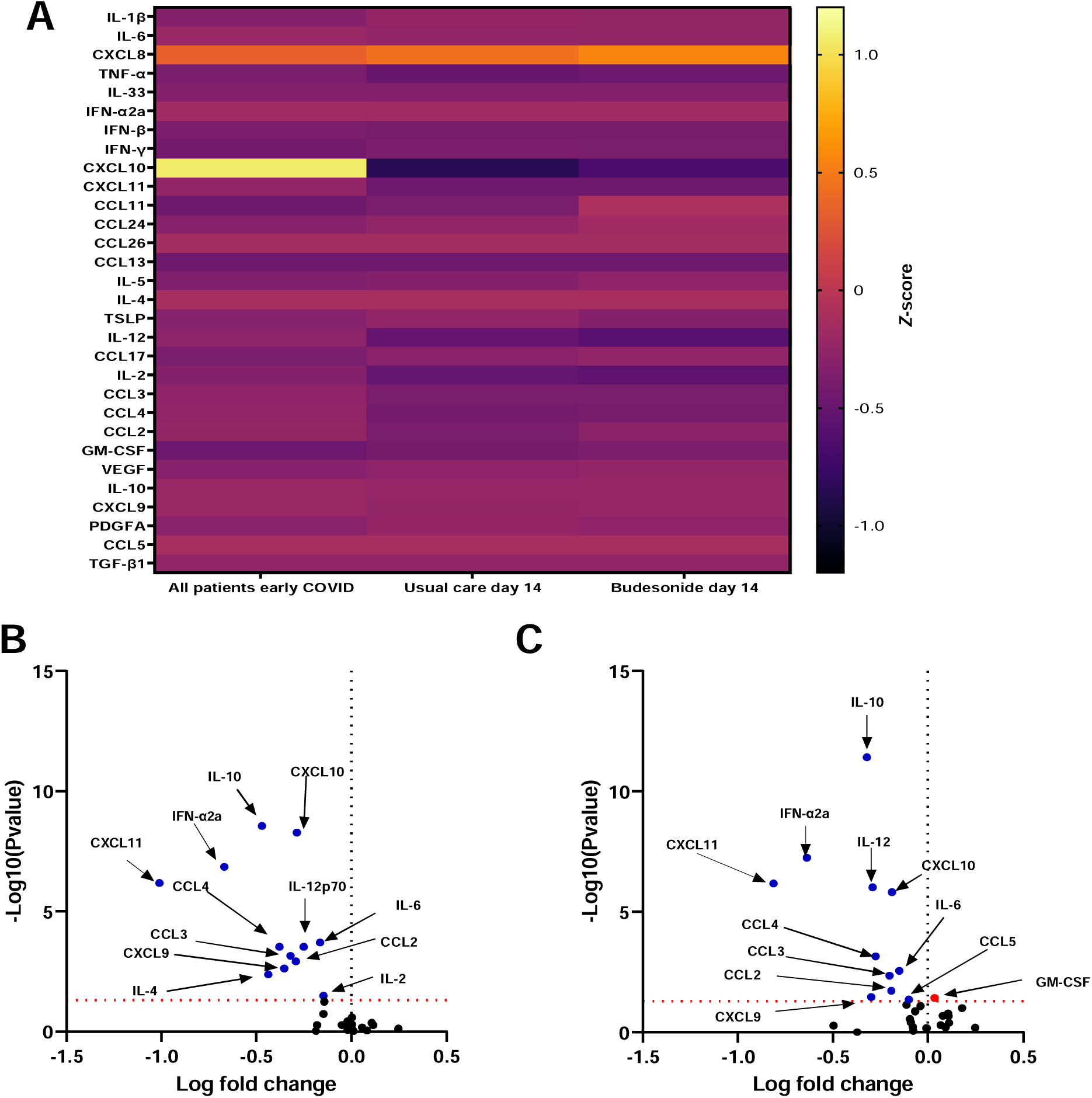
Difference in mediator levels between usual care and budesonide treatment compared to all early COVID-19 patients. A) A heatmap depicting changes in nasal mediators comparing levels in all visit 1 samples (n=140) and visit 3 in usual care arm (n=60) and budesonide arm (n=62). B) Volcano plot comparing all visit 1 samples to usual care visit 3. C) Volcano plot comparing all visit 1 samples to budesonide visit 3. ⍰= upregulated. ⍰= downregulated. Data were analysed using Mann-Whitney t-test.

**Supplementary figure S4.**
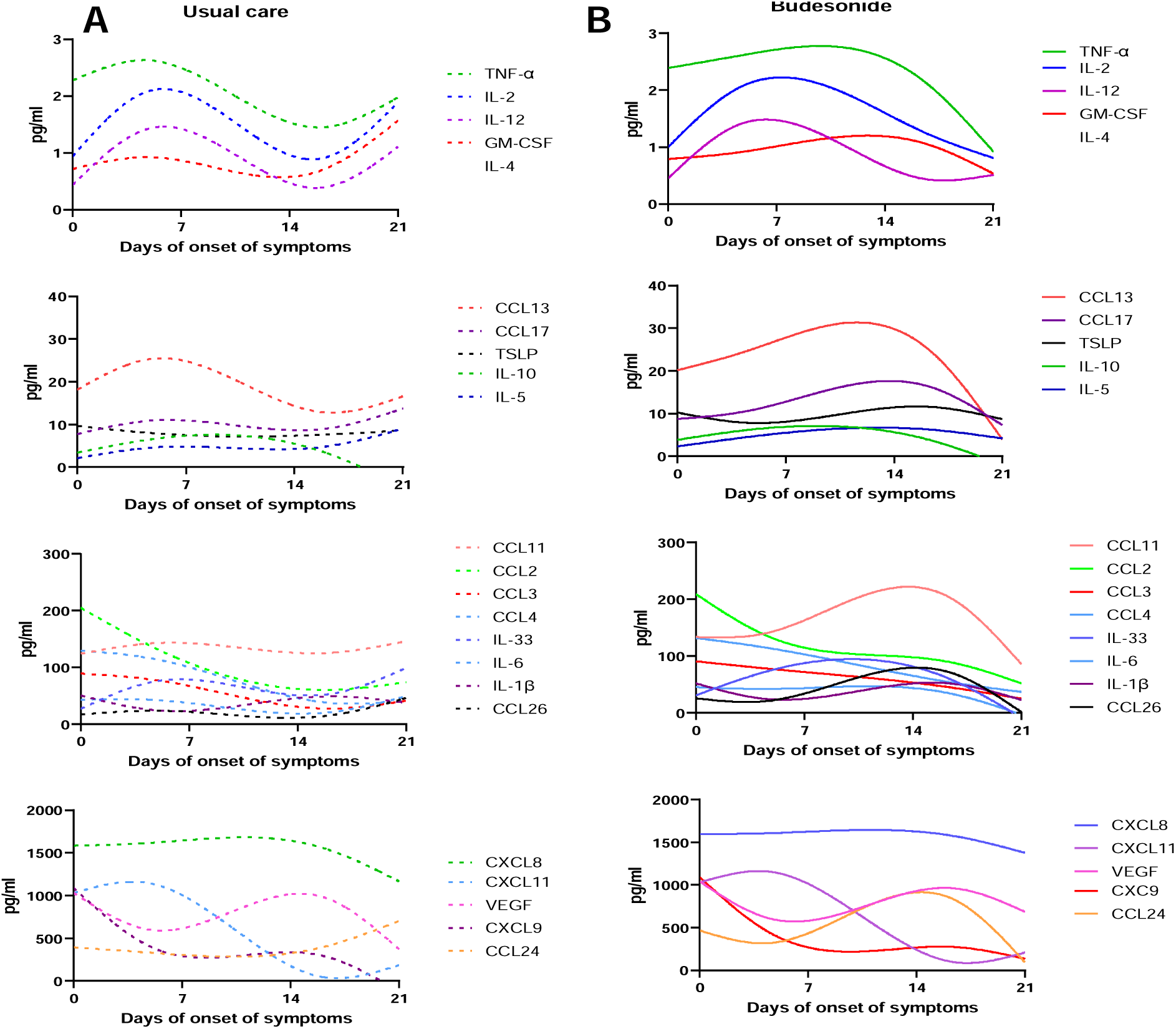
Spline analysis of nasal mucosal mediator levels from first symptom onset. Longitudinal analysis of mediator profiles in the A) usual care (n=60) and B) budesonide arm (n=62) displayed as representative best fit curves by smoothed spline analysis.

**Supplementary figure S5.**
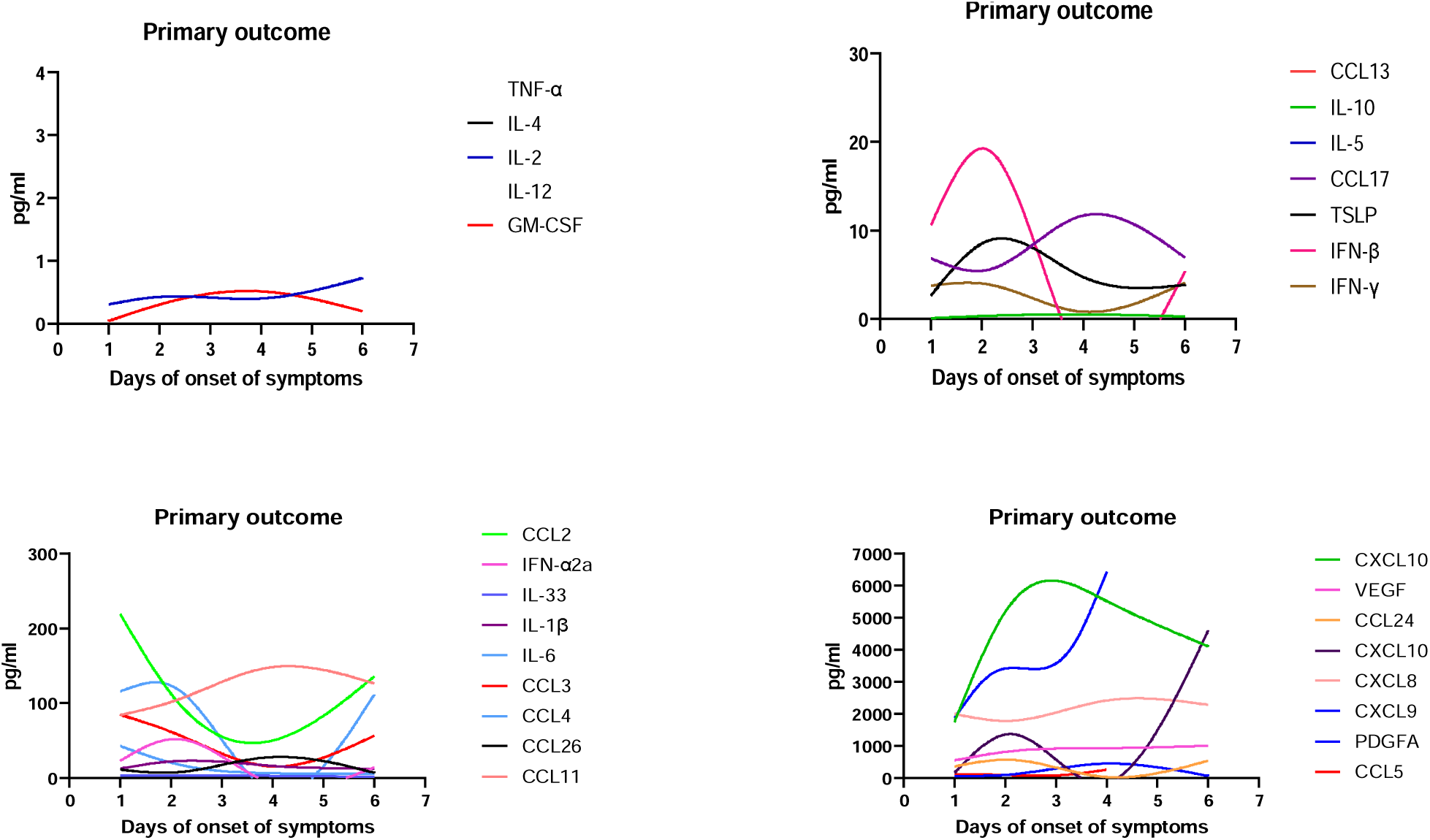
Spline analysis of nasal mucosal mediator levels from first symptom onset in participants who reached the primary outcome (n=11).

**Supplementary figure S6.**
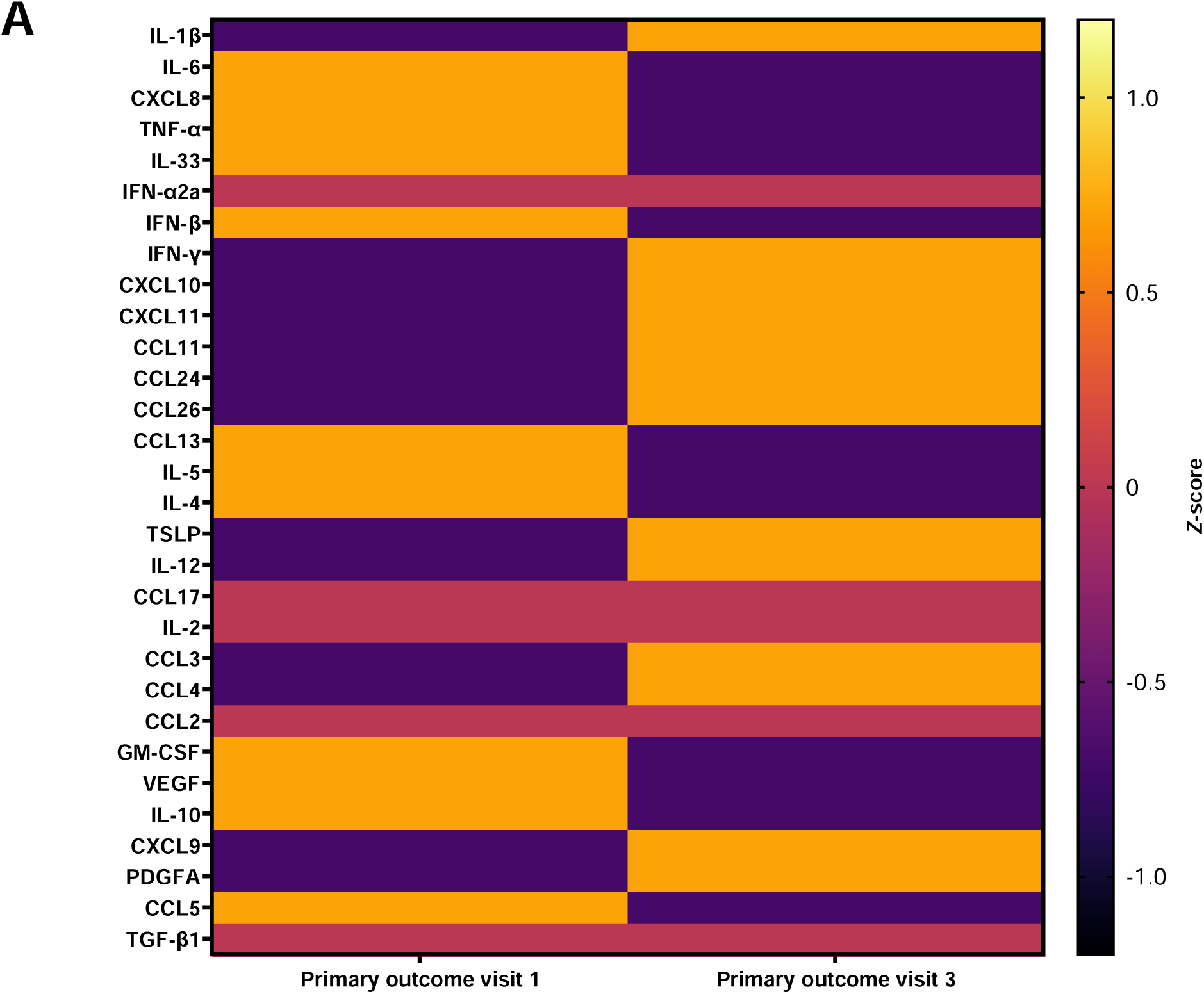
Heatmap for one patient who developed severe COVID-19 infection and met the study primary outcome who gave two nasal samples.

**Supplementary figure S7.**
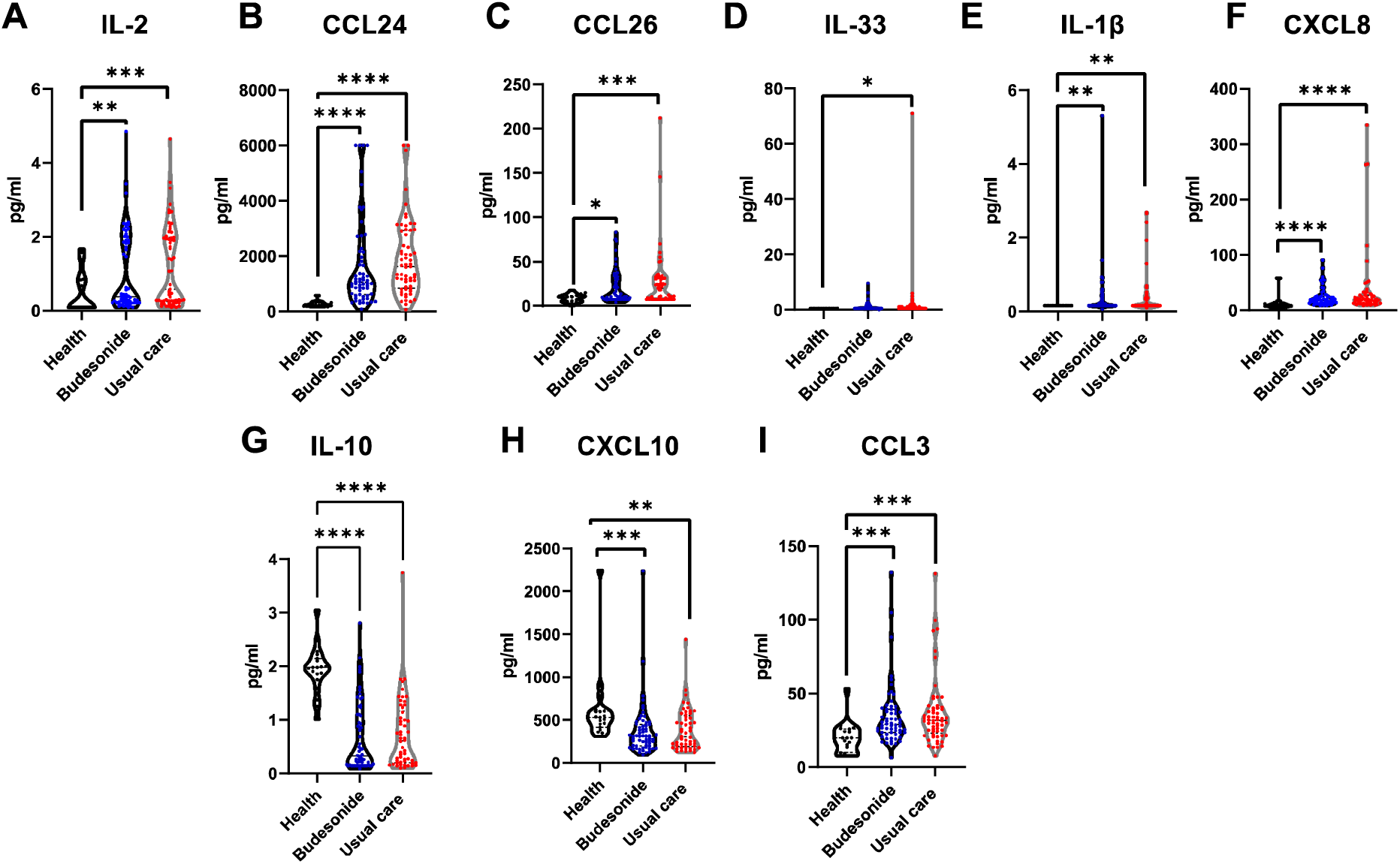
Additional systemic mediators which remain altered 28-35 days after COVID-19 infection. A-I) Violin plots comparing mediator levels in the serum of healthy individuals (n=19), those in the usual care arm (n=60) and budesonide arm (n=62) of the study at 28 days post recruitment. Data were analysed by Kruskal-Wallis with post-hoc Dunn’s test. * p<0.05, ** p<0.01, ***p<0.001, ****p<0.0001.

## Extended Data Supplementary Tables

S1 Demographic table of healthy controls

|  | Healthy nasal samples<br>(n=20) | Healthy serum samples<br>(n=19) |
| --- | --- | --- |
| Age, years | 37 (24-56) | 29 (26-49) |
| Sex |  |  |
| *Female* | 16 (80%) | 14 (74%) |
| *Male* | 4 (20%) | 5 (26%) |
| Ethnicity |  |  |
| *White* | 18 (90%) | 14 (74%) |
| *Non-white* | 2 (10%) | 5 (26%) |
| Smoking history | 16 Non-smoker<br>4 Ex-smokers | 16 Non-smoker<br>2 Ex-smokers<br>1 current |

**Supplementary Table 2.**
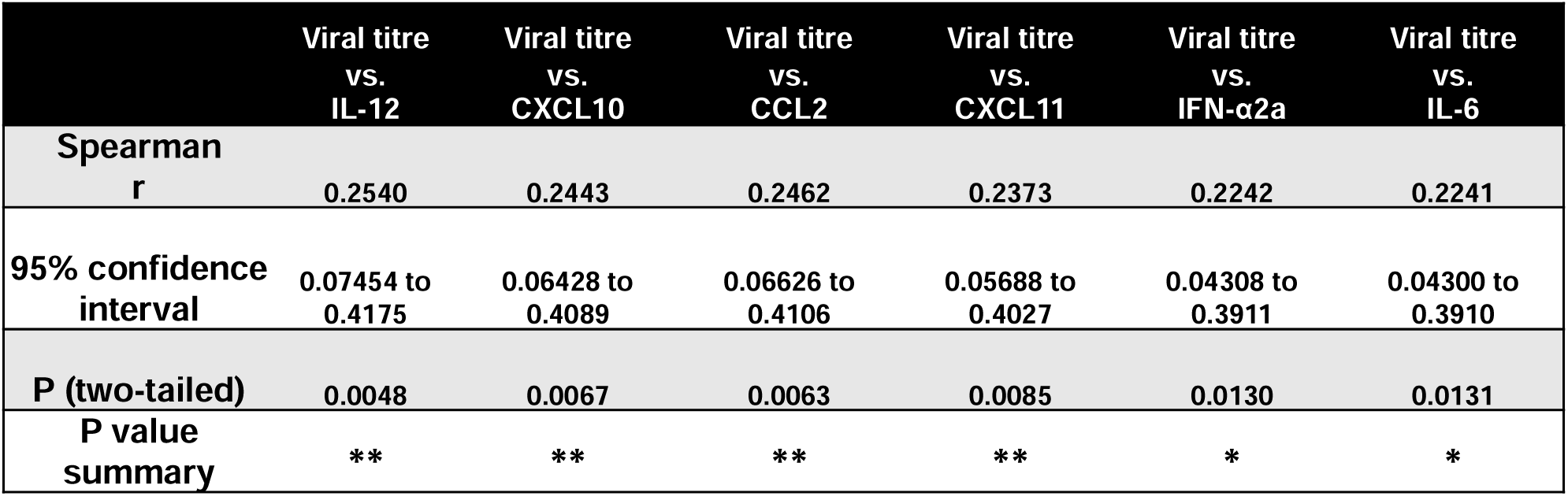
Correlation between nasal mediators and viral load. Two tailed Spearman rank correlations with 95% confidence intervals between viral load and individual mediator levels at visit 1 for those that gave both a viral swab and nasosorption sample (n=122). A). Only positively correlated mediated depicted. **= p<0.01, *= p<0.05.

| | Viral titre<br>vs.<br>IL-12 | Viral titre<br>vs.<br>CXCL10 | Viral titre<br>vs.<br>CCL2 | Viral titre<br>vs.<br>CXCL11 | Viral titre<br>vs.<br>IFN-α2a | Viral titre<br>vs.<br>IL-6 |
| --- | --- | --- | --- | --- | --- | --- |
| **Spearman**<br>**r** | 0.2540 | 0.2443 | 0.2462 | 0.2373 | 0.2242 | 0.2241 |
| **95% confidence**<br>**interval** | 0.07454 to<br>0.4175 | 0.06428 to<br>0.4089 | 0.06626 to<br>0.4106 | 0.05688 to<br>0.4027 | 0.04308 to<br>0.3911 | 0.04300 to<br>0.3910 |
| **P (two-tailed)** | 0.0048 | 0.0067 | 0.0063 | 0.0085 | 0.0130 | 0.0131 |
| **P value**<br>**summary** | **\*\*** | **\*\*** | **\*\*** | **\*\*** | **\*** | **\*** |

## Notes

### Author Declarations

The National Health Service/Health Research Authority UK Fulham London Research Ethics Committee gave ethical approval for this work (20/HRA/2531)

